# Racial disparity in distant recurrence-free survival in localized breast cancer patients: A pooled analysis of NSABP trials

**DOI:** 10.1101/2021.12.15.21267850

**Authors:** Gina Kim, Jessica M. Pastoriza, Jiyue Qin, Juan Lin, George S. Karagiannis, John S. Condeelis, Greg Yothers, Stewart Anderson, Thomas Julian, David Entenberg, Thomas E. Rohan, Xiaonan Xue, Joseph A. Sparano, Maja H. Oktay

## Abstract

**Background:** Black race is associated with worse outcome in patients with breast cancer. We evaluated distant relapse-free survival (DRFS) between Black and White women with localized breast cancer who participated in NCI-sponsored clinical trials.

**Methods:** We analyzed pooled data from eight National Surgical Adjuvant Breast and Bowel Project (NSABP) trials including 9,702 women with localized breast cancer treated with adjuvant chemotherapy (AC, n=7,485) or neoadjuvant chemotherapy (NAC, n=2,217), who self-reported as Black (n=1,070) or White (n=8,632). The association between race and DRFS was analyzed using log-rank tests and multivariate Cox regression.

**Results:** After adjustment for covariates including age, tumor size, nodal status, body mass index and taxane use, and treatment (AC vs. NAC), Black race was associated with an inferior DRFS in ER-positive (HR 1.24 [95% CI 1.05-1.46], p=0.01), but not in ER-negative disease (HR 0.97 [95% CI 0.83-1.14], p=0.73), and significant interaction between race and ER status was observed (p=0.03). There was no racial disparity in DRFS among patients with pathologic complete response (pCR) (Log-rank p =0.8). For patients without pCR, black race was associated with worse DRFS in ER-positive (HR 1.67 [95% CI 1.14-2.45], p=0.01), but not in ER-negative disease (HR 0.91 [95% CI 0.65-1.28], p=0.59).

**Conclusion:** Black race was associated with significantly inferior DRFS in ER-positive localized breast cancer treated with AC or NAC, but not in ER-negative disease. In the NAC group, racial disparity was also observed in patients with residual ER-positive breast cancer at surgery, but not in those who had a pathologic complete response.

**Lay Summary:** Black women with breast cancer have worse outcome compared to White women. We investigated if this held true in the context of clinical trials which provide controlled treatment setting. Black women with cancer containing estrogen receptors (ER) had worse outcome than White women. If breast cancers did not contain ER there was no racial disparity in outcome. Moreover, we observed racial disparity in women who received chemotherapy before their cancer was removed, but only if they had cancer with ER and residual disease upon completion of treatment. If cancer disappeared upon pre-surgical chemotherapy, there was no racial disparity.

## Background

Breast cancer is the second most commonly diagnosed cancer and the second leading cause of cancer death among U.S. women ^1,2^. Black race or African American (AA) ethnicity has been associated with worse outcome in both women and men with localized and advanced breast cancer^3-5^. This disparity has been attributed to patient- and tumor-specific factors such as more advanced disease at presentation ^6^, higher rates of obesity ^7^, and higher rates of unacceptable side effects to chemotherapy ^8,9^, as well as differences in tumor microenvironment (TME) and systemic inflammatory signatures ^10^. Other factors contributing to racial disparities include differences in socioeconomic status and limitations in access to care ^11^. Studying race and ethnicity-dependent outcome has been challenging due to heterogeneity within racial and ethnic groups and difficulty in controlling for social determinants of health.

Clinical trials provide an opportunity to control for some of the social determinants of health, such as access to care, medical comorbidities, as well as variations in treatment and adherence to therapy. However, Blacks and Hispanics are consistently underrepresented in phase III oncologic clinical trials, including trials sponsored by the U.S. National Cancer Institute (NCI) ^12,13^.

Efforts have been made to overcome this limitation by pooling data from NCI-sponsored adjuvant trials in localized breast cancer. For example, a pooled analysis of four trials (B-13, 14, 19, and 20) coordinated by the National Surgical Adjuvant Breast and Bowel Project (NSABP) including 5,444 patients with localized breast cancer that completed accrual prior to 1988 found that there was no racial disparity in cancer-specific outcomes ^14^. On the other hand, in an analysis of 35 phase III trials coordinated by the Southwestern Oncology Group (SWOG), including 19,547 patients with a variety of solid tumors and hematologic malignancies, there was a racial disparity, even after controlling for clinical prognostic factors, income, and education, among those with breast, ovarian, or prostate cancer ^15^. Similarly, Black race was associated with a worse outcome in those with hormone receptor (HR)-positive, HER2-negative disease after controlling for covariates including obesity in a trial including 4,817 women coordinated by the Eastern Cooperative Oncology Group (ECOG) ^7^. Moreover, a recent analysis of another NCI-sponsored trial (TAILORx) that included 10,273 women demonstrated a higher distant recurrence rate in Black compared to White participants with HR-positive HER2-negative disease despite similar 21-gene assay recurrence scores^16^.

Neoadjuvant chemotherapy (NAC) has emerged as an effective means to downstage locally advanced disease and to allow for breast conservation and less axillary surgery ^17^. Although single-institution, retrospective studies have demonstrated a worse distant recurrence free survival (DRFS) in Blacks compared to Whites treated with NAC ^18,19^, this has been previously investigated in only one prospective clinical trial, which showed a trend towards worse DRFS in Blacks compared to Whites, though not statistically significant^20^.

Given the discrepant results of AC trial analyses and lack of NAC studies, we aimed to evaluate racial disparity in DRFS using pooled analysis of 8 NSABP trials including 9,702 women with localized breast cancer receiving NAC or AC regimens including doxorubicin and cyclophosphamide with or without a taxane; this analysis did not include 4 trials reported in a prior NSABP analysis^14^. NSABP is an NCI-sponsored clinical trials cooperative group that has research sites at nearly 1,000 cancer treatment centers in the United States, Canada, Puerto Rico, Australia, and Ireland.

## Methods

### Study/patient selection

A pooled analysis of eight NSABP trials was performed (Appendix 1), including five trials of only adjuvant chemotherapy (B-15, B-22, B-23, B-28, B-30), one trial including both adjuvant and neoadjuvant chemotherapy arms (B-18 arm 1 and arm 2), and two trials including only neoadjuvant chemotherapy (B-27 arm 1 and arm 2, B-40 arm 1A); patients enrolled in the B-27 3^rd^ arm including both AC and NAC were not included in this analysis. The accrual dates for the trials are included in Supplemental Table 1S. These eight trials were selected by reviewing treatment with doxorubicin, cyclophosphamide with or without a taxane for localized invasive cancer. Of the trials included in our analyses, onlyB-40 had information regarding HER2 expression available. Therefore, we could not perform subgroup analysis according to HER2 expression status. Additionally, B-36 was excluded as patients were treated with trastuzumab at the discretion of treating physician. Of the remaining trials, there were 14 involving doxorubicin and cyclophosphamide. To minimize the effects of treatment variations, we only included the trials in which patients received doxorubicin 60mg/m^2^ IV and cyclophosphamide 600 mg/m^2^ IV with or without a taxane every 3 weeks (n=9). B-16 was excluded due to its limited eligibility criteria. Lastly, B-29 (arm 3) was excluded because it included <8% Black participants. AC treatment group was defined as patients who received surgery first, followed by chemotherapy. NAC treatment group was defined as those who received all chemotherapy prior to surgery (B-27 arm 3 was excluded due to receipt of taxane after surgery). Of the 10,522 patients in these trials, only patients of Black (n=1,070, 11.0%) or White (n=8,632, 89.0%) race were included in the analyses. Race was self-reported, and ethnicity was not considered (i.e., White race encompasses both Hispanic and non-Hispanic Whites).

**Table 1.** Patient characteristics, entire cohort. ǂPathologic tumor size and node status for adjuvant group; clinical (pre-treatment) tumor size and node status for neoadjuvant group **Obesity defined as BMI ≥ 30kg/m^2^

|  | **Full Sample(N=9702)** | **White(N=8632)** | **Black(N=1070)** | **P Value** |
| --- | --- | --- | --- | --- |
| **Age,Mean(SD)** | 49.4(10) | 49.7(10) | 47.6(10) | <.0001 |
| **AC/NAC,Count(%)** |  |  |  | 0.0097 |
| **AC** | 7485(77.1) | 6693(77.5) | 792(74) |  |
| **NAC** | 2217(22.9) | 1939(22.5) | 278(26) |  |
| **ER Status,Count(%)** |  |  |  | <.0001 |
| **Negative** | 3436(35.4) | 2932(34) | 504(47.1) |  |
| **Positive** | 4942(50.9) | 4551(52.7) | 391(36.5) |  |
| **Unknown** | 1324(13.6) | 1149(13.3) | 175(16.4) |  |
| **Node Status\*,Count(%)** |  |  |  | 0.0007 |
| **Negative** | 2703(27.9) | 2359(27.3) | 344(32.1) |  |
| **Positive** | 6930(71.4) | 6216(72) | 714(66.7) |  |
| **Unknown** | 69(0.7) | 57(0.7) | 12(1.1) |  |
| **Tumor Size,Count(%)** |  |  |  | 0.0003 |
| **<2 cm** | 3265(33.7) | 2959(34.3) | 306(28.6) |  |
| **$\geq 2$ cm** | 6205(64) | 5476(63.4) | 729(68.1) |  |
| **Unknown** | 232(2.4) | 197(2.3) | 35(3.3) |  |
| **Obese,Count(%)** |  |  |  | <.0001 |
| **Non-obese** | 6845(70.6) | 6293(72.9) | 552(51.6) |  |
| **Obese\*\*** | 2855(29.4) | 2337(27.1) | 518(48.4) |  |
| **Unknown** | 2(0) | 2(0) |  |  |
| **Taxel,Count(%)** |  |  |  | 0.3212 |
| **No Taxel** | 5803(59.8) | 5148(59.6) | 655(61.2) |  |
| **Taxel** | 3899(40.2) | 3484(40.4) | 415(38.8) |  |
| **BMI Group,Count(%)** |  |  |  | <.0001 |
| **<20** | 495(5.1) | 473(5.5) | 22(2.1) |  |
| **[20, 25)** | 3318(34.2) | 3132(36.3) | 186(17.4) |  |
| **[25,30)** | 3032(31.3) | 2688(31.1) | 344(32.1) |  |
| **$\geq 30$** | 2855(29.4) | 2337(27.1) | 518(48.4) |  |
| **Unknown** | 2(0) | 2(0) |  |  |

### Statistical analysis

Patient and tumor characteristics such as age (years), ER status (positive vs. negative), tumor size (≥2 cm vs. < 2 cm), nodal status (positive vs. negative), obesity status (obese vs. non-obese) and taxane use (yes vs. no), were compared using Wilcoxon rank sum tests for continuous variables and Pearson’s chi-squared tests for categorical variables. Obesity was defined as BMI ≥30kg/m^2^. Pathological tumor size and node status were used for AC group while clinical tumor size and node status were used for NAC group. Pathologic complete response (pCR) was defined as no residual disease in the breast and was also compared between Black and White patients in the NAC cohort.

The outcome evaluated was distant relapse-free survival (DRFS), defined as time to first distant relapse or a second primary cancer. Death prior to a distant recurrence or second primary cancer was censored. The median follow-up was 115.2 months, and we censored the follow-up beyond 210 months. Kaplan-Meier survival curves were estimated for each racial group (adjusted Kaplan-Meier curves and log-rank tests^21^ were also estimated, see Appendix 2) and log-rank tests were used to compare survival between Black and White. Multivariate Cox proportional hazard models stratified were used to compare survival between Black and White patients, while adjusting for age, ER status, tumor size, nodal status, obesity status and taxane use and treatment. Because prior studies indicated racial disparities were evident primarily in ER-positive breast cancer^7,16^, separate Cox models were used for ER-positive and ER-negative disease. A formal comparison of racial disparity between ER-positive and ER-negative disease was made by testing the interaction between race and ER status. Within the NAC cohort, separate analyses were also performed by pCR, because pCR pertains good long-term prognosis. Since prior studies indicated racial disparity in ER positive breast cancer^7,16^, separate analyses for patients with ER+ and ER-disease were performed in patients with residual cancer after NAC.

The proportionality of the Cox models was examined based upon Schoenfeld residuals^22,23^, and no violation was identified.

Statistical significance was specified a priori as p<0.05 and the p-values reported are two-sided. All analyses were conducted using SAS 9.4 (SAS Institute Inc., Cary, NC 2014).

This study was reviewed by the Albert Einstein College of Medicine Institutional Review Board and was deemed exempt status (IRB# 2018-8959).

## Results

### Patient characteristics

There were 9,702 patients included in our analysis, all women, of whom 1,070 (11.0%) were Black and 8632 (89.0%) were White. Of note, patients characterized as “other”, and Hispanics were not included in the analysis. It was not clear from the data provided what race was included in the “other”. Likewise, patients in the Hispanic group were not categorized as Black or White. Of these, 7,485 (77.1%) were in the AC treatment group and 2,217 (22.9 %) were in the NAC treatment group (Table 1). Compared to AC treatment group, patients in the NAC treatment group had more node negative disease (68.9% vs 15.7% p<0.001) and more often had tumors ≥2cm (93.3% vs 55.3%, p<0.001). The NAC treatment group also had a higher proportion of Black patients (12.5% vs 10.6%, p=0.01). There was no difference between treatment groups in age at diagnosis, prevalence of obesity, and receipt of taxane (Table 1). The differences in tumor size and node status were attributable to the differences in enrollment criteria for the trials (Appendix 1). Furthermore, in the AC treatment group, we compared the pathologic tumor size and node status, and in the NAC treatment group, the clinical, or pre-treatment tumor size and node status.

### Clinical outcomes in the entire cohort

In the entire cohort, Black patients had a worse DRFS compared to White patients (Log-Rank p<0.001) (Figure 1A), and Black race remained significantly associated with a worse DRFS after adjusting for age, ER status, tumor size, node status, obesity, receipt of taxane, and treatment (AC vs. NAC) (HR 1.13 95% CI 1.01-1.25, p=0.03) (Figure 1B). However, multivariate analysis stratifying patients by ER status showed that Black race was significantly associated with a worse DRFS in ER-positive disease (HR 1.24 [95% CI 1.05-1.46], p=0.01) (Figure 1C), but not in ER-negative disease (HR 0.97 [95% CI 0.83-1.14], p=0.73) (Figure 1D). The Cox model including the interaction term revealed significant interaction between race and ER status (p=0.03) (Table 2S).

**Figure 1.**
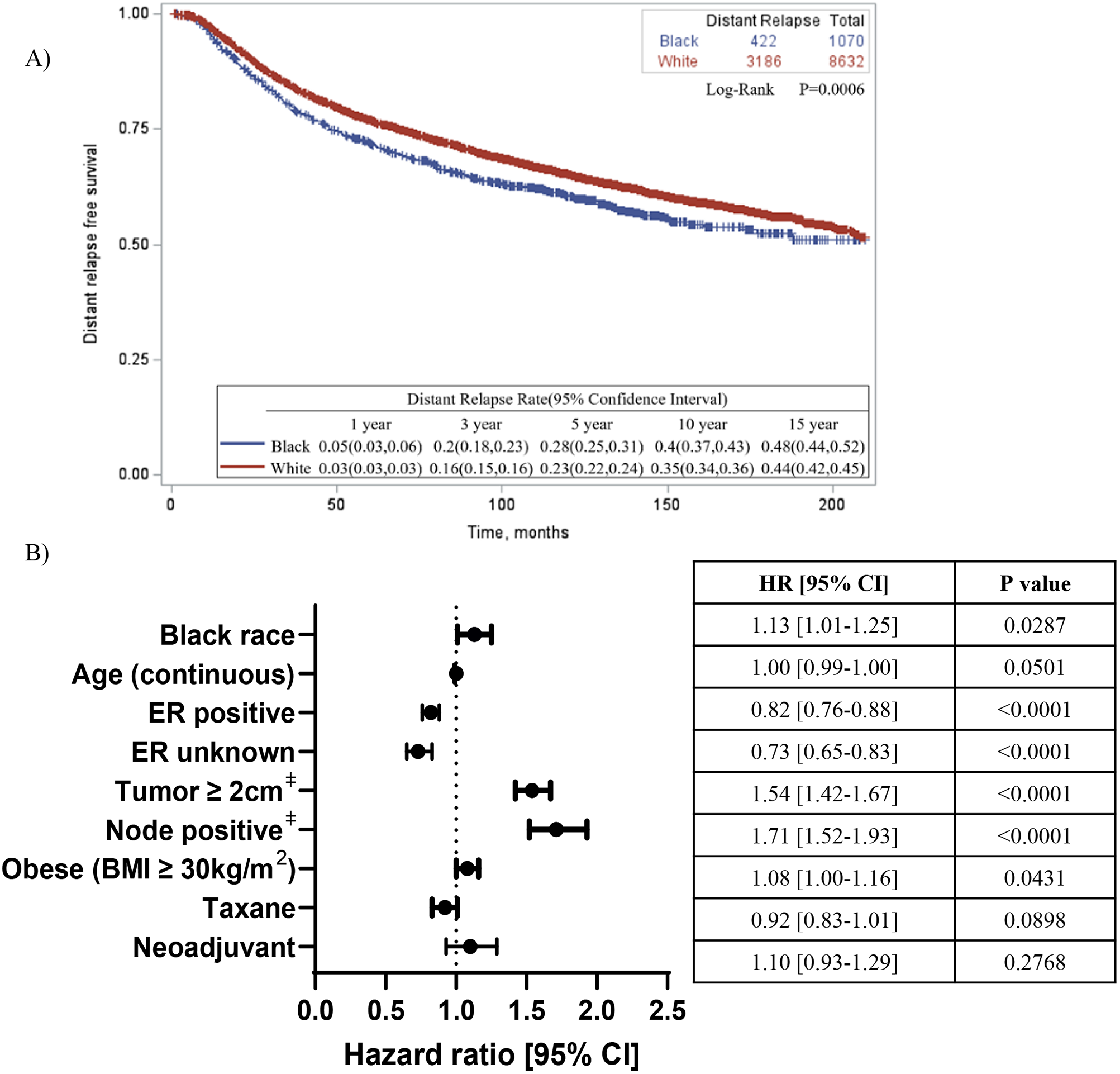
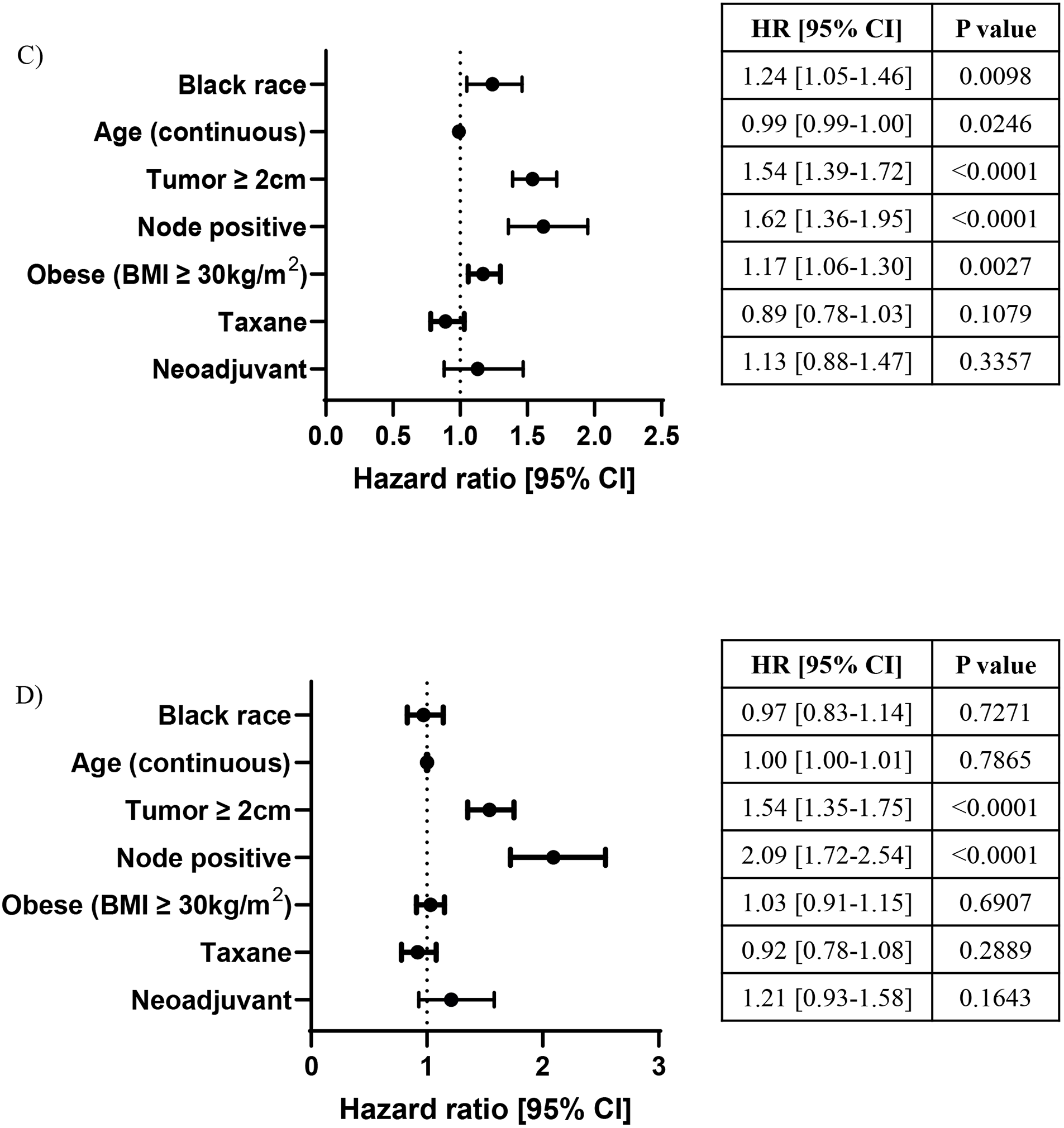
Association between race and DRFS, entire cohort. A) Kaplan-Meier curve describing distant-relapse free survival in entire cohort. Log-rank test was used to compare survival in White vs Black race patients. B) Cox-regression model adjusting for co-variates and stratified by protocol. Comparison groups: White race, ER-negative, node negative, tumor <2cm, non-obese (BMI <30kg/m^2^), no taxane, adjuvant chemotherapy. C) Cox-regression model in ER-positive disease only. D) Cox-regression model in ER-negative disease only. ‡Pathologic tumor size and node status for the adjuvant group; clinical (pre-treatment) tumor size and node status for the neoadjuvant group

Given the differences in tumor characteristics between AC (Table 3S) and NAC (Table 4S) treatment groups, we repeated the analyses stratified by use of AC or NAC. Although, Black patients had worse DRFS compared to White patients the association was not statistically significant after adjustment for covariates including obesity (Figures 1S and 2S).

### Clinical outcomes in the NAC treatment group

As pCR portends to better long-term outcome, the NAC treatment group was further stratified by pCR status. There were 394 patients (17.8%) who achieved pCR, of whom 61(15.5%) were Black and 333 (84.5%) were White. There were 1,731 (81.5%) patients who did not achieve pCR, of whom 198 (11.4%) were Black and 1,533 (88.6%) were White. pCR status was unknown in 92 (4.3%) of the patients. Among patients who achieved pCR, there was no racial disparity in DRFS (Log-Rank p=0.81) (Figure 2A). Among those who did not achieve pCR, however, Black patients had a significantly worse DRFS (Log-Rank p=0.002) (Figure 2B). However, after adjusting for covariates, Black race was not significantly associated with a worse DRFS (HR 1.23 95% CI [0.98-1.53], p=0.08) (Figure 2C).

**Figure 2.**
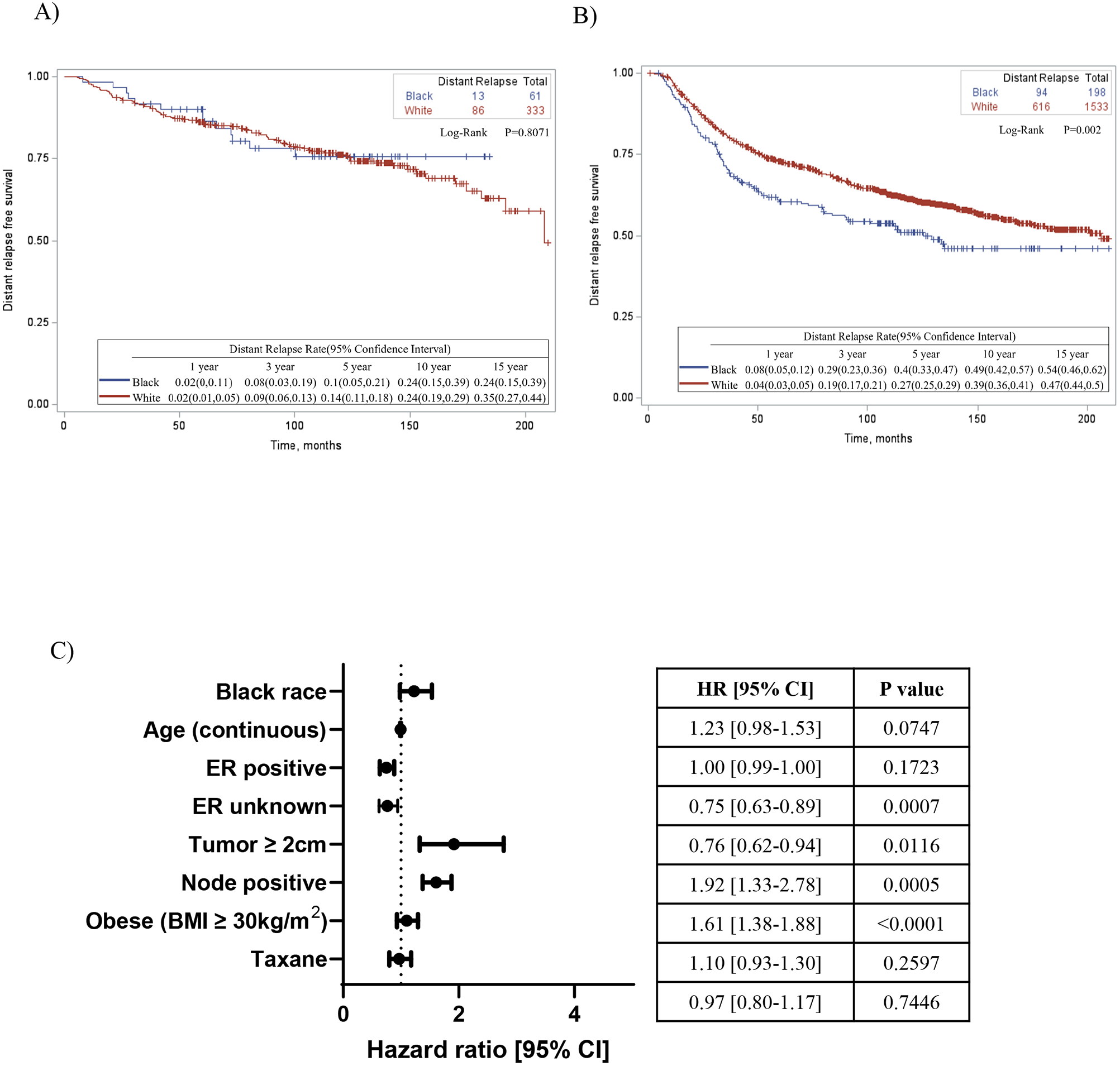
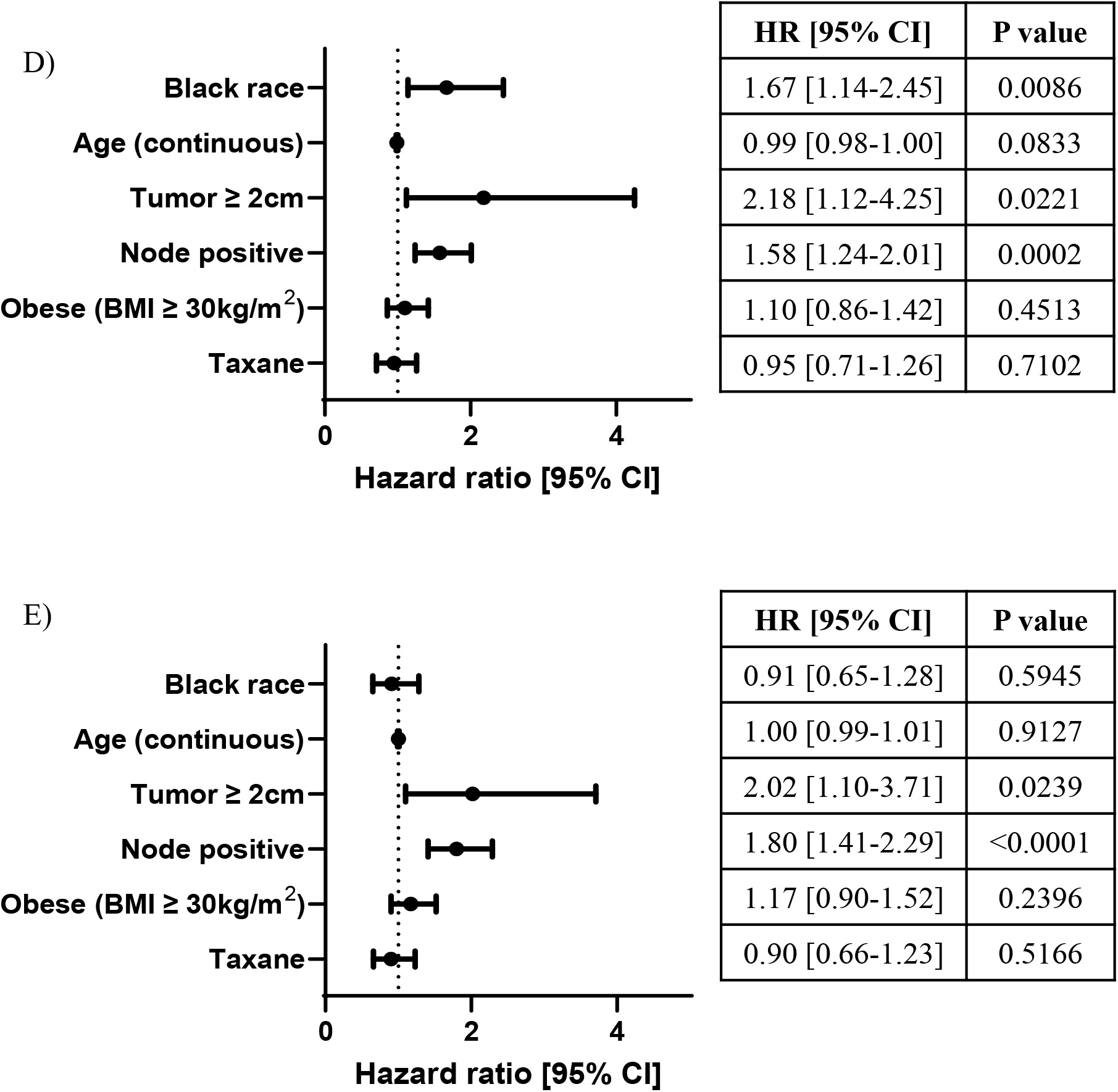
Association between race and DRFS in neoadjuvant treatment cohort, stratified by pCR. A) Kaplan-Meier curve describing distant-relapse free survival in patients who achieved pathologic complete response following neoadjuvant chemotherapy. Log-rank and Wilcoxon tests were used to compare survival in White vs Black race patients. B) Kaplan-Meier curve describing distant-relapse free survival in patients who did not achieve pathologic complete response following neoadjuvant chemotherapy. Log-rank and Wilcoxon tests were used to compare survival in White vs Black race patients. C) Cox-regression model adjusting for co-variates in no pCR only. D) Cox-regression model in ER-positive disease with no pCR. E) Cox-regression model in ER-negative disease with no pCR. Comparison groups: White race, ER-negative, node negative, tumor <2cm, non-obese (BMI <30kg/m^2^), no taxane.

Because prior studies show racial disparity in patients with ER+ disease the NAC treatment group without pCR was stratified by ER status (Figure 2D, E). In ER-positive disease, Black race was significantly associated with a worse DRFS even after adjusting for all covariates (HR 1.67 [95% CI 1.14-2.45], p=0.009) while no significant association with DRFS was found in ER-negative group (HR 0.91 [95% CI 0.65-1.28], p=0.59). Although the estimates of racial disparity were very different between the ER groups, the interaction between race and ER status was not significant (p=0.13) (Table 5S).

In summary, there was no racial disparity in DRFS among patients who achieved pCR. For patients who did not achieve pCR, black race was associated with worse DRFS in ER-positive disease, but not in ER-negative disease.

## Discussion

In this pooled analysis of data from eight prospective NCI-sponsored clinical trials, we found that after adjusting for covariates including age, tumor size, node status, obesity, receipt of taxane and treatment (AC vs. NAC), Black race was associated with a worse DRFS in ER-positive disease but not in ER-negative disease. Formal testing in the entire cohort demonstrated a significant interaction between race and ER status. Within the NAC cohort, there was no racial disparity in DRFS among patients who achieved pCR. For patients who did not achieve pCR, black race was only associated with worse DRFS in ER-positive disease, but not in ER-negative disease. However, formal testing did not reveal a significant interaction between race and ER status, likely due to a limited sample size.

Utilizing data from patients who qualify for participation in national NCI-sponsored clinical trials minimizes some of the potential factors contributing to racial disparities noted in population-based studies, including access to care and significant comorbidities. Furthermore, utilizing clinical trial data ensured administration of and adherence to a pre-determined treatment regimen. However, the social determinants of health were not considered in these trials. The patient accrual took place more than twenty years ago, when there was less known about how these parameters may affect the long-term outcome of some patients.

The findings of this study are in line with the existing literature that Black women receiving chemotherapy for localized breast cancer have an inferior DRFS ^14,24-28^. Consistent with prior reports on racial disparity in ER+ disease^7,16^, Black race was significantly associated with higher distant recurrence rates in ER-positive disease when adjusted for other prognostic covariates in the overall cohort. In addition, we showed here that Black race was associated with worse DRFS in patients with ER+ residual tumors following neoadjuvant therapy. The decision to stratify according to ER status and pCR was also based on parameters affecting long term outcomes in patients. ER positive, compared to ER negative disease tends to have a weaker response to chemotherapy, and pCR is associated with a better long-term outcome. Further research is needed to better understand whether the observed disparity is due to adherence to endocrine therapy, and/or biological, environmental and/or social determinants of health.

Consistent with prior reports, we did not observe substantial racial disparities in pCR rates after NAC^29-32^. In fact, Black women in our analysis had somewhat higher pCR rates than Whites (21.9% vs. 17.2%, p=0.0051), which is likely due to a higher prevalence of basal-like or triple negative subtype in Blacks ^33,34^ that is more chemosensitive ^35-39^. Due to limitations in the dataset, we were not able to evaluate molecular subtypes in this study directly, although the higher prevalence of ER-negative disease among Blacks would suggest that Black women in our group have a higher prevalence of basal-like subtype in line with the existing literature^40^.

Strengths of this study include the large number of patients included in the analysis, the long follow-up duration, inclusion of subjects who had access to care and absence of comorbidities that would preclude optimal adjuvant systemic chemotherapy and endocrine therapy, and the use of standard state-of-the-art chemotherapy regimens in controlled clinical settings. Limitations include use of self-reported race, absence of information regarding social determinants of health, and absence of information on treatment administration and adherence. Race is a social construct, and self-identified race has an imperfect correlation with geographical ancestry as determined by genetic studies ^41-43^. Furthermore, both Black and White races are heterogeneous populations, and there is emerging evidence that presentation and outcomes within Black race vary widely depending on additional ethnic variables such as country of origin ^44-46^. This intra-racial heterogeneity merits further exploration in future analyses and prospective collection of genetic ancestry may be a step towards personalized prognosis and treatment options. Regarding social determinants of health, patient eligibility for enrollment to clinical trials controls for some but not all of these factors. One study found that after adjusting for poverty and Medicaid enrollment, race was not in and of itself a risk factor for worse survival ^47^. However, a pooled analysis of two ECOG clinical trials found that insurance and neighborhood SES did not affect outcomes ^48^.

In conclusion, in this pooled analysis of eight NSABP trials including 9,702 women with localized breast cancer treated with adjuvant or neoadjuvant systemic chemotherapy including cyclophosphamide and doxorubicin without a taxane, Black women with ER-positive disease had a worse DRFS compared to White women, especially in those with residual cancer following NAC. Further research is needed to fully understand the mechanisms behind this disparity.

## Data Availability

All data produced in the present study are available upon reasonable request to the authors

## Notes

### Author contributions

GK, JMP, GSK, DE, XX, MHO—conceptualization; GK, JQ, SA, TJ,GY —data curation; JQ, JL, SA, GY, XX—Formal analysis & methodology; GK, JMP— writing – original draft; All authors—Writing – review & editing

### Prior presentations

This work was presented at *American Association for Cancer Research Annual Meeting* on April 10, 2021 (virtual meeting).

## Acknowledgements

We would like to acknowledge *NRG Oncology/NSABP* for providing us the data that was used for this study.

## References

1. DeSantis CE, Ma J, Gaudet MM, et al: Breast cancer statistics, 2019. CA Cancer J Clin 69:438–451, 2019

2. Siegel RL, Miller KD, Fuchs HE, et al: Cancer Statistics, 2021. CA Cancer J Clin 71:7–33, 2021

3. DeSantis CE, Miller KD, Goding Sauer A, et al: Cancer statistics for African Americans, 2019. CA Cancer J Clin 69:211–233, 2019

4. Shin JY, Kachnic LA, Hirsch AE: The impact of race in male breast cancer treatment and outcome in the United States: a population-based analysis of 4,279 patients. Int J Breast Cancer 2014:685842, 2014

5. Sineshaw HM, Freedman RA, Ward EM, et al: Black/White Disparities in Receipt of Treatment and Survival Among Men With Early-Stage Breast Cancer. J Clin Oncol 33:2337–44, 2015

6. Joslyn SA, West MM: Racial differences in breast carcinoma survival. Cancer 88:114–23, 2000

7. Sparano JA, Wang M, Zhao F, et al: Race and hormone receptor-positive breast cancer outcomes in a randomized chemotherapy trial. J Natl Cancer Inst 104:406–14, 2012

8. Hershman D, McBride R, Jacobson JS, et al: Racial disparities in treatment and survival among women with early-stage breast cancer. J Clin Oncol 23:6639–46, 2005

9. Schneider BP, Li L, Radovich M, et al: Genome-Wide Association Studies for Taxane-Induced Peripheral Neuropathy in ECOG-5103 and ECOG-1199. Clin Cancer Res 21:5082–5091, 2015

10. Kim G, Pastoriza JM, Condeelis JS, et al: The Contribution of Race to Breast Tumor Microenvironment Composition and Disease Progression. Front Oncol 10:1022, 2020

11. Tammemagi CM, Nerenz D, Neslund-Dudas C, et al: Comorbidity and survival disparities among black and white patients with breast cancer. JAMA 294:1765–72, 2005

12. Gopishetty S, Kota V, Guddati AK: Age and race distribution in patients in phase III oncology clinical trials. Am J Transl Res 12:5977–5983, 2020

13. Loree JM, Anand S, Dasari A, et al: Disparity of Race Reporting and Representation in Clinical Trials Leading to Cancer Drug Approvals From 2008 to 2018. JAMA Oncol:e191870, 2019

14. Dignam JJ: Efficacy of systemic adjuvant therapy for breast cancer in African-American and Caucasian women. J Natl Cancer Inst Monogr:36-43, 2001

15. Albain K GR SJ, Makower DF, Pritchard KI, Hayes DF, Geyer JR.CE, Dees EC, Goetz MP, Olson Jr.: JA, Lively T, Badve SS, Saphner TJ, Wagner LI, Whelan TJ, Ellis MJ, Paik S, Wood WC, Ravdin PM, Keane MM, Gomez HL, Reddy PS, Goggins TF, MAyer IA, Brufsky AM, Toppmeyer DL, Kaklamani VG, Berenberg JL, Abrams J, Sledge, Jr. Gw.: Race, ethnicity and clinical outcomes in hormone receptor-positive, HER2-negative, node negative breast cancer: results from the TAILORx trial. Cancer Research, 2018

16. Albain KS, Gray RJ, Makower DF, et al: Race, Ethnicity, and Clinical Outcomes in Hormone Receptor-Positive, HER2-Negative, Node-Negative Breast Cancer in the Randomized TAILORx Trial. J Natl Cancer Inst 113:390–399, 2021

17. Fisher B, Brown A, Mamounas E, et al: Effect of preoperative chemotherapy on local-regional disease in women with operable breast cancer: findings from National Surgical Adjuvant Breast and Bowel Project B-18. J Clin Oncol 15:2483–93, 1997

18. Howard-McNatt M, Lawrence J, Melin SA, et al: Race and recurrence in women who undergo neoadjuvant chemotherapy for breast cancer. Am J Surg 205:397–401, 2013

19. Pastoriza JM, Karagiannis GS, Lin J, et al: Black race and distant recurrence after neoadjuvant or adjuvant chemotherapy in breast cancer. Clin Exp Metastasis 35:613–623, 2018

20. Woodward WA, Huang EH, McNeese MD, et al: African-American race is associated with a poorer overall survival rate for breast cancer patients treated with mastectomy and doxorubicin-based chemotherapy. Cancer 107:2662–8, 2006

21. Xie J, Liu C: Adjusted Kaplan-Meier estimator and log-rank test with inverse probability of treatment weighting for survival data. Stat Med 24:3089–110, 2005

22. Grambsch PaTT: Proportional hazards tests and diagnostics based on weighted residuals.. Biometrika 81:515–526, 1994

23. Schoenfeld D: Partical residuals for the proportional hazards regression model.. Biometrika 69:239–241, 1982

24. Jatoi I, Anderson WF, Rao SR, et al: Breast cancer trends among black and white women in the United States. J Clin Oncol 23:7836–41, 2005

25. Kabat GC, Ginsberg M, Sparano JA, et al: Risk of Recurrence and Mortality in a Multi-Ethnic Breast Cancer Population. J Racial Ethn Health Disparities 4:1181–1188, 2017

26. Menashe I, Anderson WF, Jatoi I, et al: Underlying causes of the black-white racial disparity in breast cancer mortality: a population-based analysis. J Natl Cancer Inst 101:993–1000, 2009

27. Newman LA, Kaljee LM: Health Disparities and Triple-Negative Breast Cancer in African American Women: A Review. JAMA Surg 152:485–493, 2017

28. Wojcik BE, Spinks MK, Optenberg SA: Breast carcinoma survival analysis for African American and white women in an equal-access health care system. Cancer 82:1310–8, 1998

29. Mancino AT, Rubio IT, Henry-Tillman R, et al: Racial differences in breast cancer survival: the effect of residual disease. J Surg Res 100:161–5, 2001

30. Chavez-Macgregor M, Litton J, Chen H, et al: Pathologic complete response in breast cancer patients receiving anthracycline- and taxane-based neoadjuvant chemotherapy: evaluating the effect of race/ethnicity. Cancer 116:4168–77, 2010

31. Tichy JR, Deal AM, Anders CK, et al: Race, response to chemotherapy, and outcome within clinical breast cancer subtypes. Breast Cancer Res Treat 150:667–74, 2015

32. Warner ET, Ballman KV, Strand C, et al: Impact of race, ethnicity, and BMI on achievement of pathologic complete response following neoadjuvant chemotherapy for breast cancer: a pooled analysis of four prospective Alliance clinical trials (A151426). Breast Cancer Res Treat 159:109–18, 2016

33. Howlader N, Altekruse SF, Li CI, et al: US incidence of breast cancer subtypes defined by joint hormone receptor and HER2 status. J Natl Cancer Inst 106, 2014

34. Warner ET, Tamimi RM, Hughes ME, et al: Racial and Ethnic Differences in Breast Cancer Survival: Mediating Effect of Tumor Characteristics and Sociodemographic and Treatment Factors. J Clin Oncol 33:2254–61, 2015

35. Biswas T, Efird JT, Prasad S, et al: The survival benefit of neoadjuvant chemotherapy and pCR among patients with advanced stage triple negative breast cancer. Oncotarget 8:112712–112719, 2017

36. Carey LA, Dees EC, Sawyer L, et al: The triple negative paradox: primary tumor chemosensitivity of breast cancer subtypes. Clin Cancer Res 13:2329–34, 2007

37. Liedtke C, Mazouni C, Hess KR, et al: Response to neoadjuvant therapy and long-term survival in patients with triple-negative breast cancer. J Clin Oncol 26:1275–81, 2008

38. Rouzier R, Perou CM, Symmans WF, et al: Breast cancer molecular subtypes respond differently to preoperative chemotherapy. Clin Cancer Res 11:5678–85, 2005

39. von Minckwitz G, Untch M, Blohmer JU, et al: Definition and impact of pathologic complete response on prognosis after neoadjuvant chemotherapy in various intrinsic breast cancer subtypes. J Clin Oncol 30:1796–804, 2012

40. Allott EH, Geradts J, Cohen SM, et al: Frequency of breast cancer subtypes among African American women in the AMBER consortium. Breast Cancer Res 20:12, 2018

41. Kaseniit KE, Haque IS, Goldberg JD, et al: Genetic ancestry analysis on >93,000 individuals undergoing expanded carrier screening reveals limitations of ethnicity-based medical guidelines. Genet Med 22:1694–1702, 2020

42. Lee YL, Teitelbaum S, Wolff MS, et al: Comparing genetic ancestry and self-reported race/ethnicity in a multiethnic population in New York City. J Genet 89:417–23, 2010

43. Yaeger R, Avila-Bront A, Abdul K, et al: Comparing genetic ancestry and self-described race in african americans born in the United States and in Africa. Cancer Epidemiol Biomarkers Prev 17:1329–38, 2008

44. Barreto-Coelho P, Cerbon D, Schlumbrecht M, et al: Differences in breast cancer outcomes amongst Black US-born and Caribbean-born immigrants. Breast Cancer Res Treat 178:433–440, 2019

45. Camacho-Rivera M, Kalwar T, Sanmugarajah J, et al: Heterogeneity of breast cancer clinical characteristics and outcome in US black women--effect of place of birth. Breast J 20:489–95, 2014

46. Sung H, DeSantis CE, Fedewa SA, et al: Breast cancer subtypes among Eastern-African-born black women and other black women in the United States. Cancer 125:3401–3411, 2019

47. Bradley CJ, Given CW, Roberts C: Race, socioeconomic status, and breast cancer treatment and survival. J Natl Cancer Inst 94:490–6, 2002

48. Obeng-Gyasi S, O’Neill A, Zhao F, et al: Impact of insurance and neighborhood socioeconomic status on clinical outcomes in therapeutic clinical trials for breast cancer. Cancer Med 10:45–52, 2021

