## Appendix 1 and 2, Supplement Tables 1-5, Supplement Figures 1 and 2 for "Racial disparity in distant recurrence-free survival in localized breast cancer patients: A pooled analysis of NSABP trials"

### Slide 1
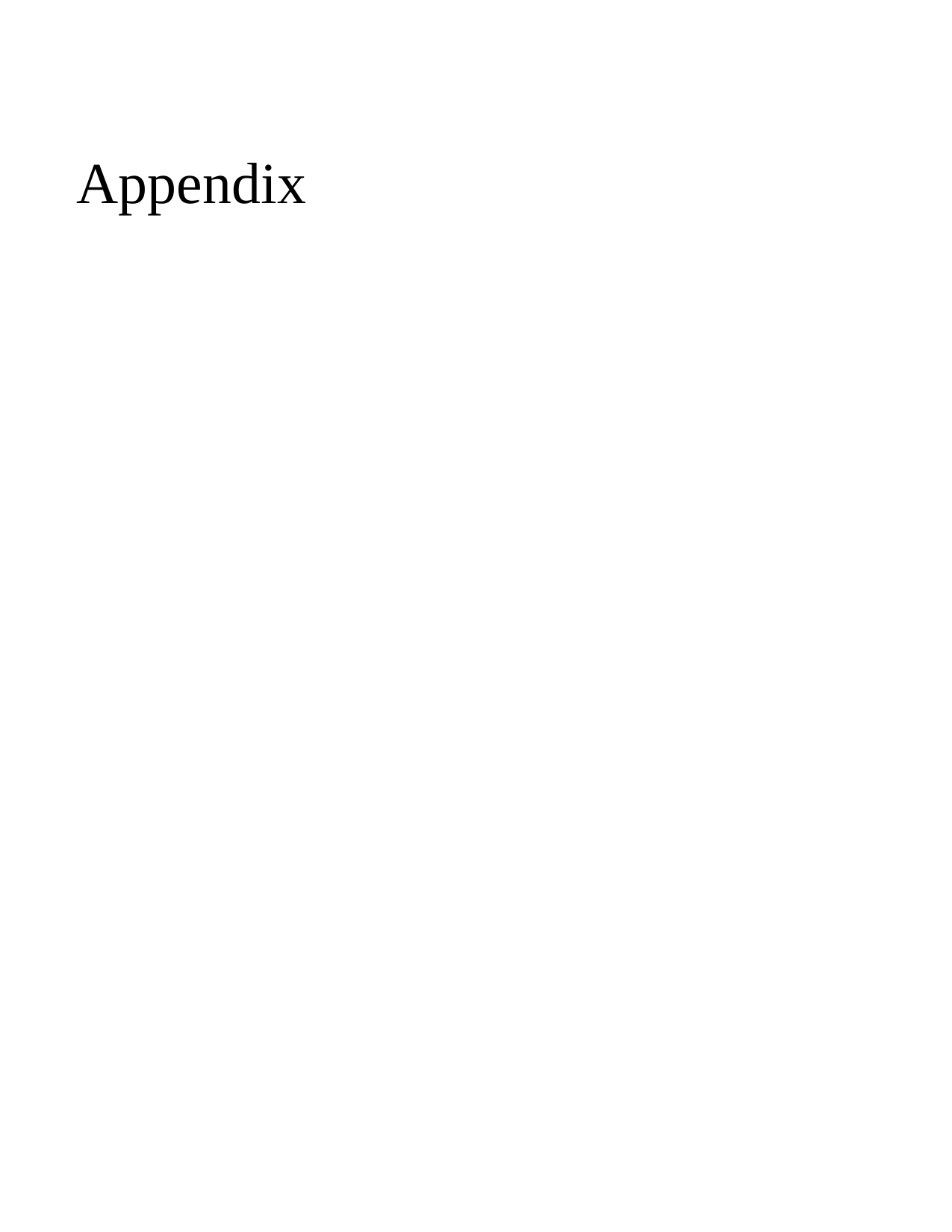

#### Appendix

### Slide 2
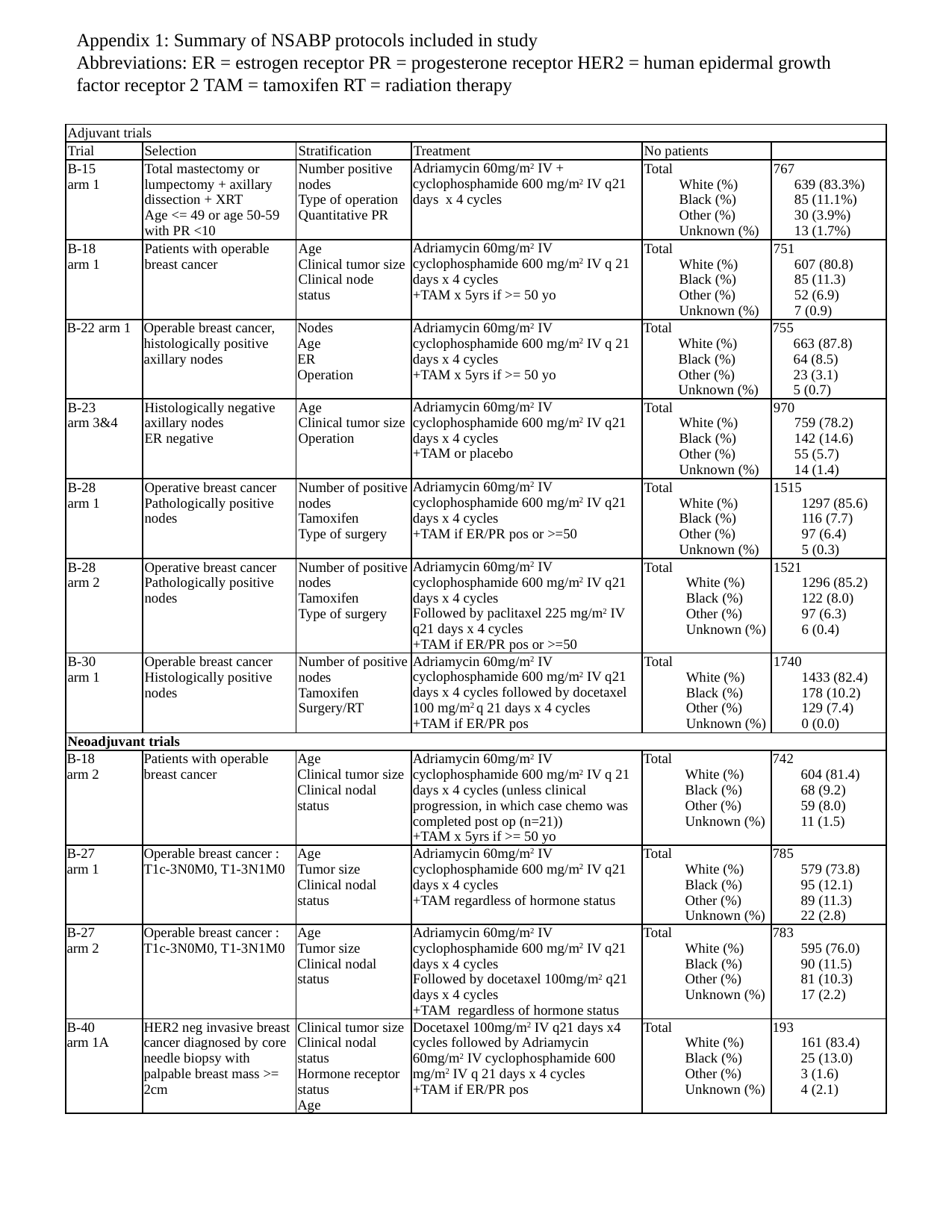

Appendix 1: Summary of NSABP protocols included in study
Abbreviations: ER = estrogen receptor PR = progesterone receptor HER2 = human epidermal growth factor receptor 2 TAM = tamoxifen RT = radiation therapy
| Adjuvant trials | | | | | |
| --- | --- | --- | --- | --- | --- |
| Trial | Selection | Stratification | Treatment | No patients | |
| B-15 arm 1 | Total mastectomy or lumpectomy + axillary dissection + XRT Age <= 49 or age 50-59 with PR <10 | Number positive nodes Type of operation Quantitative PR | Adriamycin 60mg/m^2^ IV + cyclophosphamide 600 mg/m^2^ IV q21 days x 4 cycles | Total White (%) Black (%) Other (%) Unknown (%) | 767 639 (83.3%) 85 (11.1%) 30 (3.9%) 13 (1.7%) |
| B-18 arm 1 | Patients with operable breast cancer | Age Clinical tumor size Clinical node status | Adriamycin 60mg/m^2^ IV cyclophosphamide 600 mg/m^2^ IV q 21 days x 4 cycles +TAM x 5yrs if >= 50 yo | Total White (%) Black (%) Other (%) Unknown (%) | 751 607 (80.8) 85 (11.3) 52 (6.9) 7 (0.9) |
| B-22 arm 1 | Operable breast cancer, histologically positive axillary nodes | Nodes Age ER Operation | Adriamycin 60mg/m^2^ IV cyclophosphamide 600 mg/m^2^ IV q 21 days x 4 cycles +TAM x 5yrs if >= 50 yo | Total White (%) Black (%) Other (%) Unknown (%) | 755 663 (87.8) 64 (8.5) 23 (3.1) 5 (0.7) |
| B-23 arm 3&4 | Histologically negative axillary nodes ER negative | Age Clinical tumor size Operation | Adriamycin 60mg/m^2^ IV cyclophosphamide 600 mg/m^2^ IV q21 days x 4 cycles +TAM or placebo | Total White (%) Black (%) Other (%) Unknown (%) | 970 759 (78.2) 142 (14.6) 55 (5.7) 14 (1.4) |
| B-28 arm 1 | Operative breast cancer Pathologically positive nodes | Number of positive nodes Tamoxifen Type of surgery | Adriamycin 60mg/m^2^ IV cyclophosphamide 600 mg/m^2^ IV q21 days x 4 cycles +TAM if ER/PR pos or >=50 | Total White (%) Black (%) Other (%) Unknown (%) | 1515 1297 (85.6) 116 (7.7) 97 (6.4) 5 (0.3) |
| B-28 arm 2 | Operative breast cancer Pathologically positive nodes | Number of positive nodes Tamoxifen Type of surgery | Adriamycin 60mg/m^2^ IV cyclophosphamide 600 mg/m^2^ IV q21 days x 4 cycles Followed by paclitaxel 225 mg/m^2^ IV q21 days x 4 cycles +TAM if ER/PR pos or >=50 | Total White (%) Black (%) Other (%) Unknown (%) | 1521 1296 (85.2) 122 (8.0) 97 (6.3) 6 (0.4) |
| B-30 arm 1 | Operable breast cancer Histologically positive nodes | Number of positive nodes Tamoxifen Surgery/RT | Adriamycin 60mg/m^2^ IV cyclophosphamide 600 mg/m^2^ IV q21 days x 4 cycles followed by docetaxel 100 mg/m^2^ q 21 days x 4 cycles +TAM if ER/PR pos | Total White (%) Black (%) Other (%) Unknown (%) | 1740 1433 (82.4) 178 (10.2) 129 (7.4) 0 (0.0) |
| Neoadjuvant trials | | | | | |
| B-18 arm 2 | Patients with operable breast cancer | Age Clinical tumor size Clinical nodal status | Adriamycin 60mg/m^2^ IV cyclophosphamide 600 mg/m^2^ IV q 21 days x 4 cycles (unless clinical progression, in which case chemo was completed post op (n=21)) +TAM x 5yrs if >= 50 yo | Total White (%) Black (%) Other (%) Unknown (%) | 742 604 (81.4) 68 (9.2) 59 (8.0) 11 (1.5) |
| B-27 arm 1 | Operable breast cancer : T1c-3N0M0, T1-3N1M0 | Age Tumor size Clinical nodal status | Adriamycin 60mg/m^2^ IV cyclophosphamide 600 mg/m^2^ IV q21 days x 4 cycles +TAM regardless of hormone status | Total White (%) Black (%) Other (%) Unknown (%) | 785 579 (73.8) 95 (12.1) 89 (11.3) 22 (2.8) |
| B-27 arm 2 | Operable breast cancer : T1c-3N0M0, T1-3N1M0 | Age Tumor size Clinical nodal status | Adriamycin 60mg/m^2^ IV cyclophosphamide 600 mg/m^2^ IV q21 days x 4 cycles Followed by docetaxel 100mg/m^2^ q21 days x 4 cycles +TAM regardless of hormone status | Total White (%) Black (%) Other (%) Unknown (%) | 783 595 (76.0) 90 (11.5) 81 (10.3) 17 (2.2) |
| B-40 arm 1A | HER2 neg invasive breast cancer diagnosed by core needle biopsy with palpable breast mass >= 2cm | Clinical tumor size Clinical nodal status Hormone receptor status Age | Docetaxel 100mg/m^2^ IV q21 days x4 cycles followed by Adriamycin 60mg/m^2^ IV cyclophosphamide 600 mg/m^2^ IV q 21 days x 4 cycles +TAM if ER/PR pos | Total White (%) Black (%) Other (%) Unknown (%) | 193 161 (83.4) 25 (13.0) 3 (1.6) 4 (2.1) |

### Slide 3
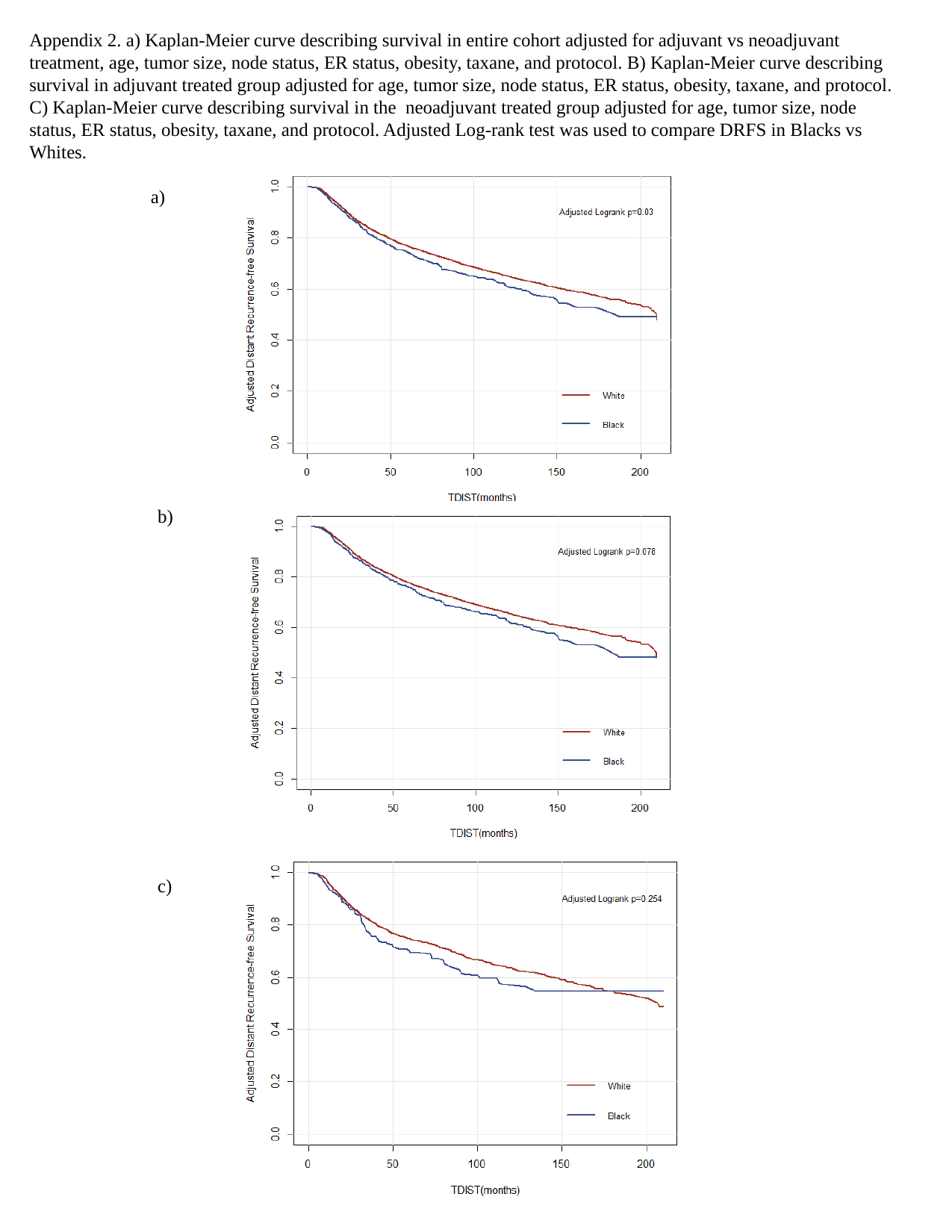

Appendix 2. a) Kaplan-Meier curve describing survival in entire cohort adjusted for adjuvant vs neoadjuvant treatment, age, tumor size, node status, ER status, obesity, taxane, and protocol. B) Kaplan-Meier curve describing survival in adjuvant treated group adjusted for age, tumor size, node status, ER status, obesity, taxane, and protocol. C) Kaplan-Meier curve describing survival in the neoadjuvant treated group adjusted for age, tumor size, node status, ER status, obesity, taxane, and protocol. Adjusted Log-rank test was used to compare DRFS in Blacks vs Whites.
a)
b)
c)

### Slide 4
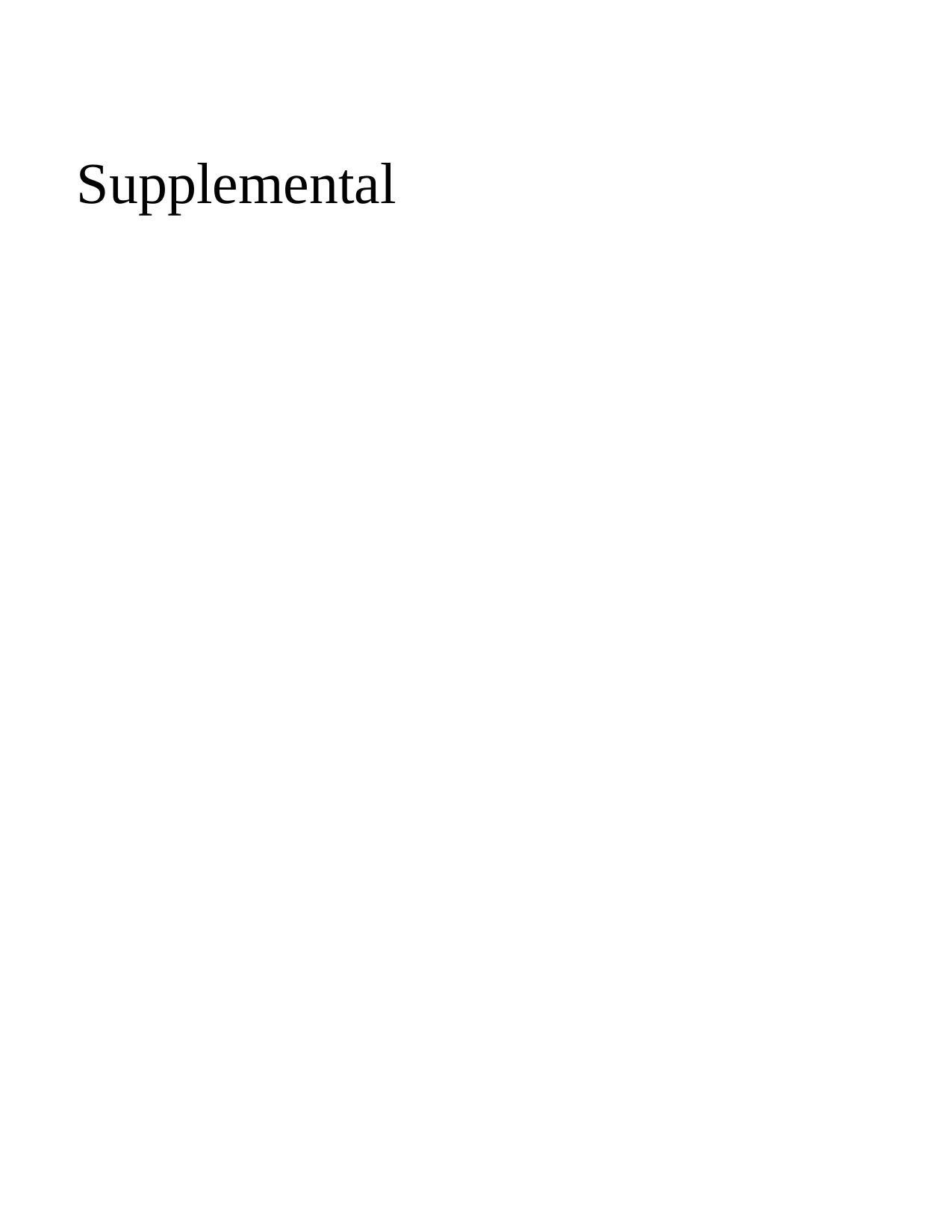

#### Supplemental

### Slide 5
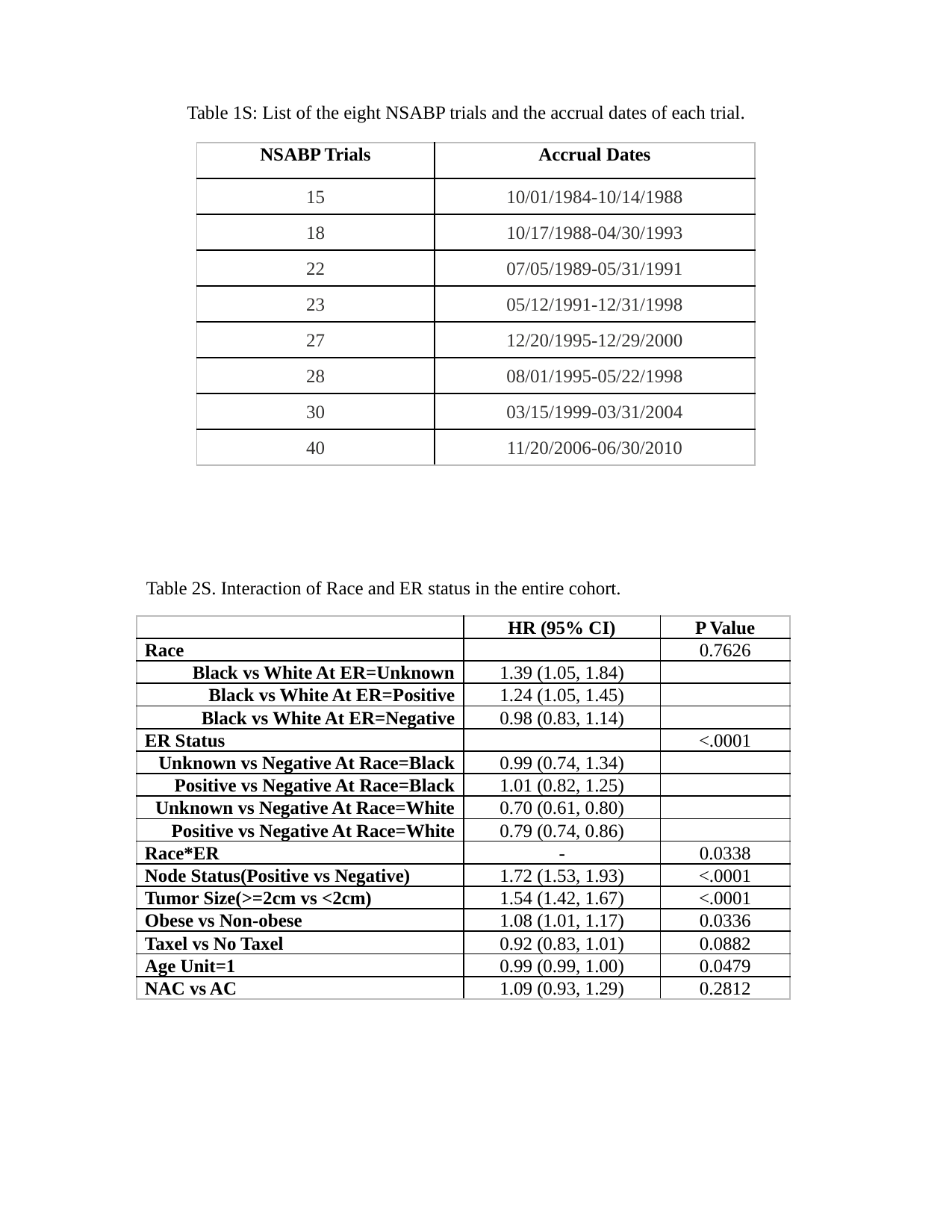

Table 1S: List of the eight NSABP trials and the accrual dates of each trial.
| NSABP Trials | Accrual Dates |
| --- | --- |
| 15 | 10/01/1984-10/14/1988 |
| 18 | 10/17/1988-04/30/1993 |
| 22 | 07/05/1989-05/31/1991 |
| 23 | 05/12/1991-12/31/1998 |
| 27 | 12/20/1995-12/29/2000 |
| 28 | 08/01/1995-05/22/1998 |
| 30 | 03/15/1999-03/31/2004 |
| 40 | 11/20/2006-06/30/2010 |
Table 2S. Interaction of Race and ER status in the entire cohort.
| | HR (95% CI) | P Value |
| --- | --- | --- |
| Race | | 0.7626 |
| Black vs White At ER=Unknown | 1.39 (1.05, 1.84) | |
| Black vs White At ER=Positive | 1.24 (1.05, 1.45) | |
| Black vs White At ER=Negative | 0.98 (0.83, 1.14) | |
| ER Status | | <.0001 |
| Unknown vs Negative At Race=Black | 0.99 (0.74, 1.34) | |
| Positive vs Negative At Race=Black | 1.01 (0.82, 1.25) | |
| Unknown vs Negative At Race=White | 0.70 (0.61, 0.80) | |
| Positive vs Negative At Race=White | 0.79 (0.74, 0.86) | |
| Race\*ER | - | 0.0338 |
| Node Status(Positive vs Negative) | 1.72 (1.53, 1.93) | <.0001 |
| Tumor Size(>=2cm vs <2cm) | 1.54 (1.42, 1.67) | <.0001 |
| Obese vs Non-obese | 1.08 (1.01, 1.17) | 0.0336 |
| Taxel vs No Taxel | 0.92 (0.83, 1.01) | 0.0882 |
| Age Unit=1 | 0.99 (0.99, 1.00) | 0.0479 |
| NAC vs AC | 1.09 (0.93, 1.29) | 0.2812 |

### Slide 6
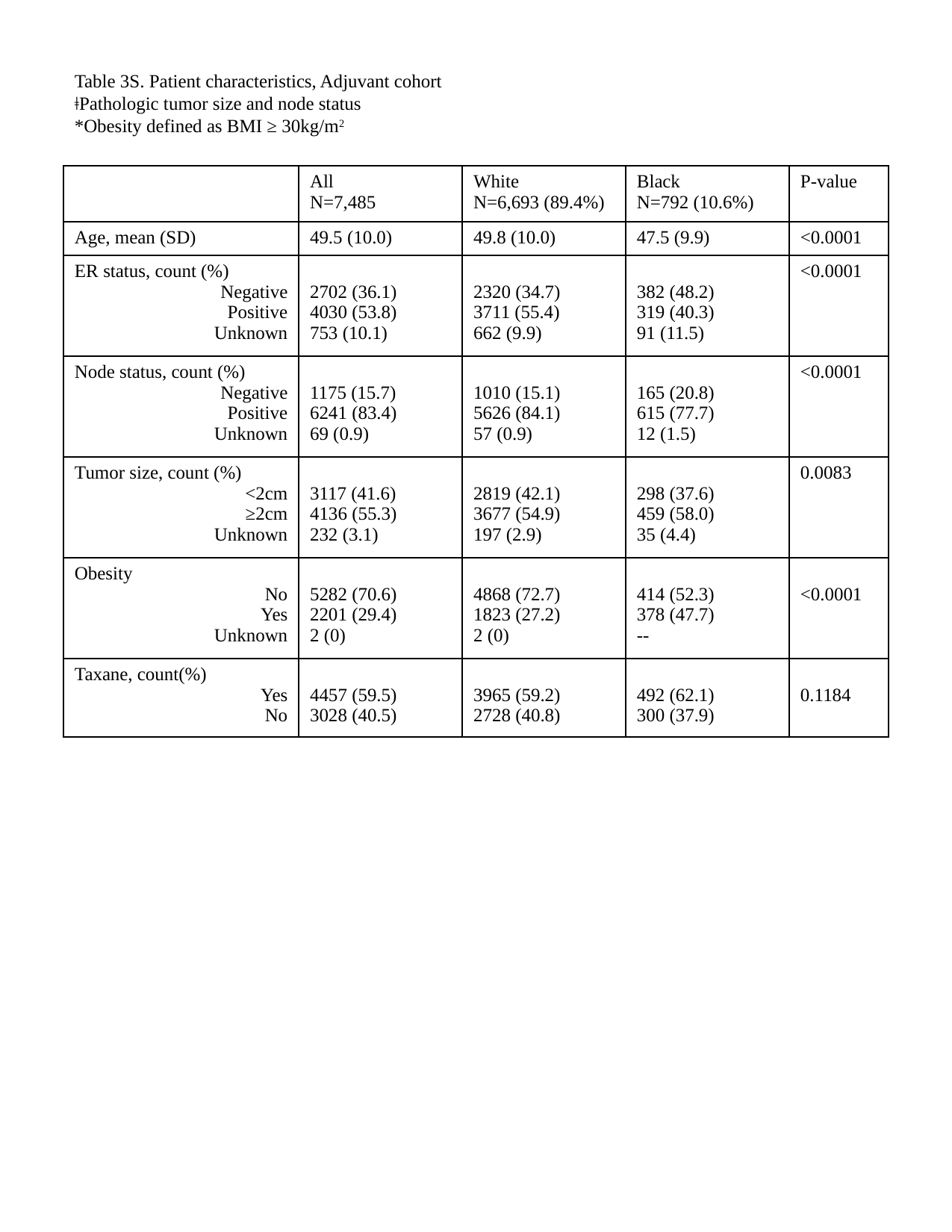

Table 3S. Patient characteristics, Adjuvant cohort
ǂPathologic tumor size and node status
*Obesity defined as BMI ≥ 30kg/m^2^
| | All N=7,485 | White N=6,693 (89.4%) | Black N=792 (10.6%) | P-value |
| --- | --- | --- | --- | --- |
| Age, mean (SD) | 49.5 (10.0) | 49.8 (10.0) | 47.5 (9.9) | <0.0001 |
| ER status, count (%) Negative Positive Unknown | 2702 (36.1) 4030 (53.8) 753 (10.1) | 2320 (34.7) 3711 (55.4) 662 (9.9) | 382 (48.2) 319 (40.3) 91 (11.5) | <0.0001 |
| Node status, count (%) Negative Positive Unknown | 1175 (15.7) 6241 (83.4) 69 (0.9) | 1010 (15.1) 5626 (84.1) 57 (0.9) | 165 (20.8) 615 (77.7) 12 (1.5) | <0.0001 |
| Tumor size, count (%) <2cm ≥2cm Unknown | 3117 (41.6) 4136 (55.3) 232 (3.1) | 2819 (42.1) 3677 (54.9) 197 (2.9) | 298 (37.6) 459 (58.0) 35 (4.4) | 0.0083 |
| Obesity No Yes Unknown | 5282 (70.6) 2201 (29.4) 2 (0) | 4868 (72.7) 1823 (27.2) 2 (0) | 414 (52.3) 378 (47.7) -- | <0.0001 |
| Taxane, count(%) Yes No | 4457 (59.5) 3028 (40.5) | 3965 (59.2) 2728 (40.8) | 492 (62.1) 300 (37.9) | 0.1184 |

### Slide 7
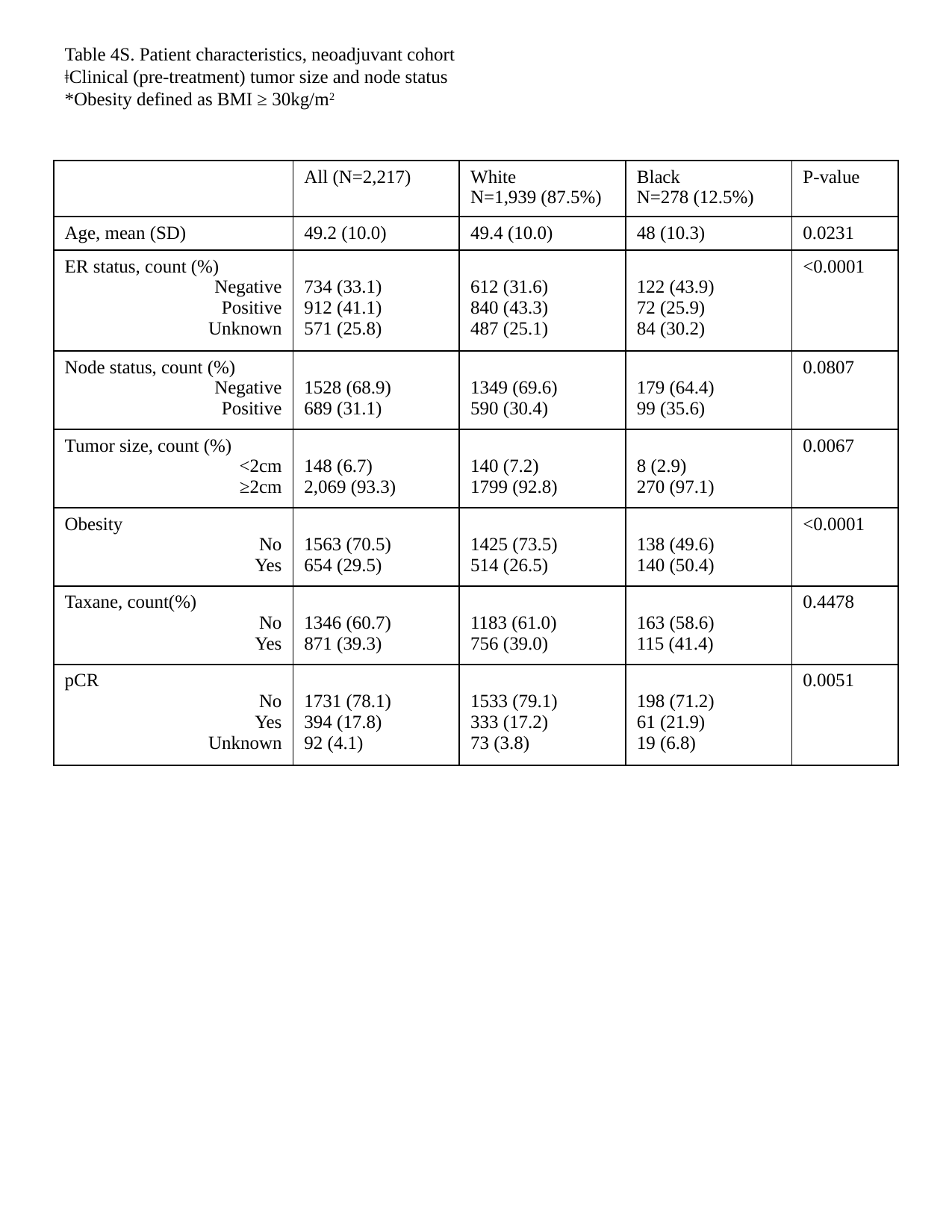

Table 4S. Patient characteristics, neoadjuvant cohort
ǂClinical (pre-treatment) tumor size and node status
*Obesity defined as BMI ≥ 30kg/m^2^
| | All (N=2,217) | White N=1,939 (87.5%) | Black N=278 (12.5%) | P-value |
| --- | --- | --- | --- | --- |
| Age, mean (SD) | 49.2 (10.0) | 49.4 (10.0) | 48 (10.3) | 0.0231 |
| ER status, count (%) Negative Positive Unknown | 734 (33.1) 912 (41.1) 571 (25.8) | 612 (31.6) 840 (43.3) 487 (25.1) | 122 (43.9) 72 (25.9) 84 (30.2) | <0.0001 |
| Node status, count (%) Negative Positive | 1528 (68.9) 689 (31.1) | 1349 (69.6) 590 (30.4) | 179 (64.4) 99 (35.6) | 0.0807 |
| Tumor size, count (%) <2cm ≥2cm | 148 (6.7) 2,069 (93.3) | 140 (7.2) 1799 (92.8) | 8 (2.9) 270 (97.1) | 0.0067 |
| Obesity No Yes | 1563 (70.5) 654 (29.5) | 1425 (73.5) 514 (26.5) | 138 (49.6) 140 (50.4) | <0.0001 |
| Taxane, count(%) No Yes | 1346 (60.7) 871 (39.3) | 1183 (61.0) 756 (39.0) | 163 (58.6) 115 (41.4) | 0.4478 |
| pCR No Yes Unknown | 1731 (78.1) 394 (17.8) 92 (4.1) | 1533 (79.1) 333 (17.2) 73 (3.8) | 198 (71.2) 61 (21.9) 19 (6.8) | 0.0051 |

### Slide 8
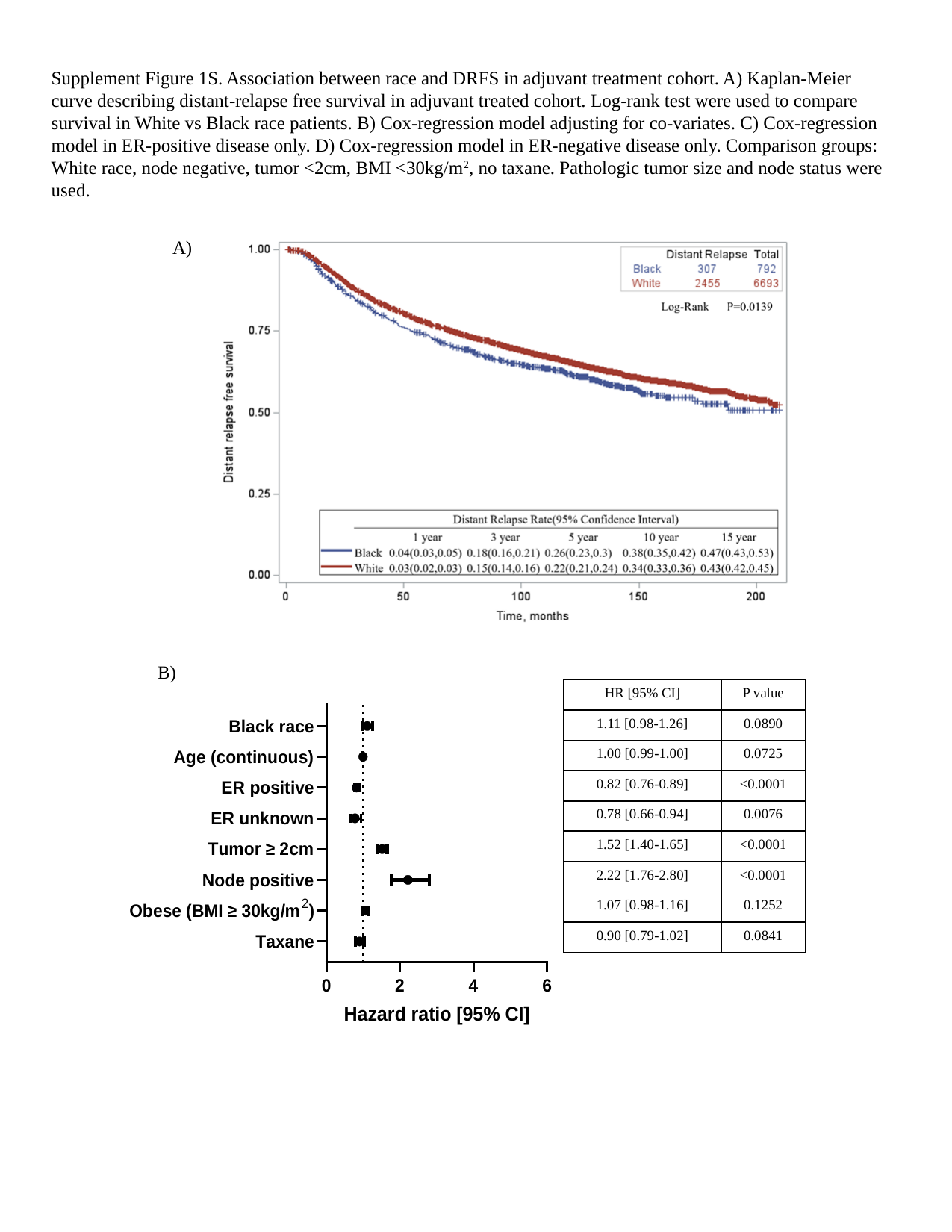

Supplement Figure 1S. Association between race and DRFS in adjuvant treatment cohort. A) Kaplan-Meier curve describing distant-relapse free survival in adjuvant treated cohort. Log-rank test were used to compare survival in White vs Black race patients. B) Cox-regression model adjusting for co-variates. C) Cox-regression model in ER-positive disease only. D) Cox-regression model in ER-negative disease only. Comparison groups: White race, node negative, tumor <2cm, BMI <30kg/m^2^, no taxane. Pathologic tumor size and node status were used.
A)
B)
| HR [95% CI] | P value |
| --- | --- |
| 1.11 [0.98-1.26] | 0.0890 |
| 1.00 [0.99-1.00] | 0.0725 |
| 0.82 [0.76-0.89] | <0.0001 |
| 0.78 [0.66-0.94] | 0.0076 |
| 1.52 [1.40-1.65] | <0.0001 |
| 2.22 [1.76-2.80] | <0.0001 |
| 1.07 [0.98-1.16] | 0.1252 |
| 0.90 [0.79-1.02] | 0.0841 |

### Slide 9
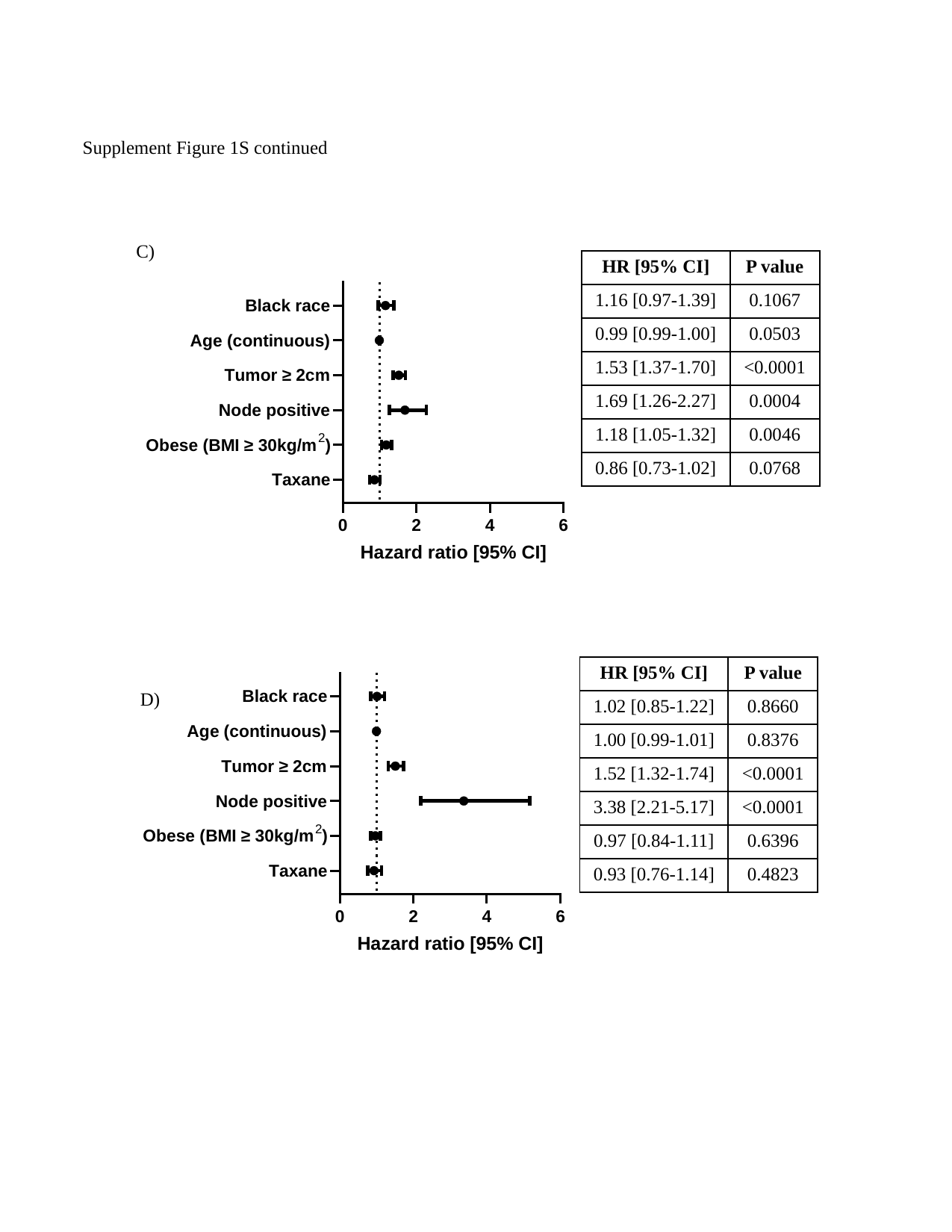

Supplement Figure 1S continued
C)
| HR [95% CI] | P value |
| --- | --- |
| 1.16 [0.97-1.39] | 0.1067 |
| 0.99 [0.99-1.00] | 0.0503 |
| 1.53 [1.37-1.70] | <0.0001 |
| 1.69 [1.26-2.27] | 0.0004 |
| 1.18 [1.05-1.32] | 0.0046 |
| 0.86 [0.73-1.02] | 0.0768 |
| HR [95% CI] | P value |
| --- | --- |
| 1.02 [0.85-1.22] | 0.8660 |
| 1.00 [0.99-1.01] | 0.8376 |
| 1.52 [1.32-1.74] | <0.0001 |
| 3.38 [2.21-5.17] | <0.0001 |
| 0.97 [0.84-1.11] | 0.6396 |
| 0.93 [0.76-1.14] | 0.4823 |
D)

### Slide 10
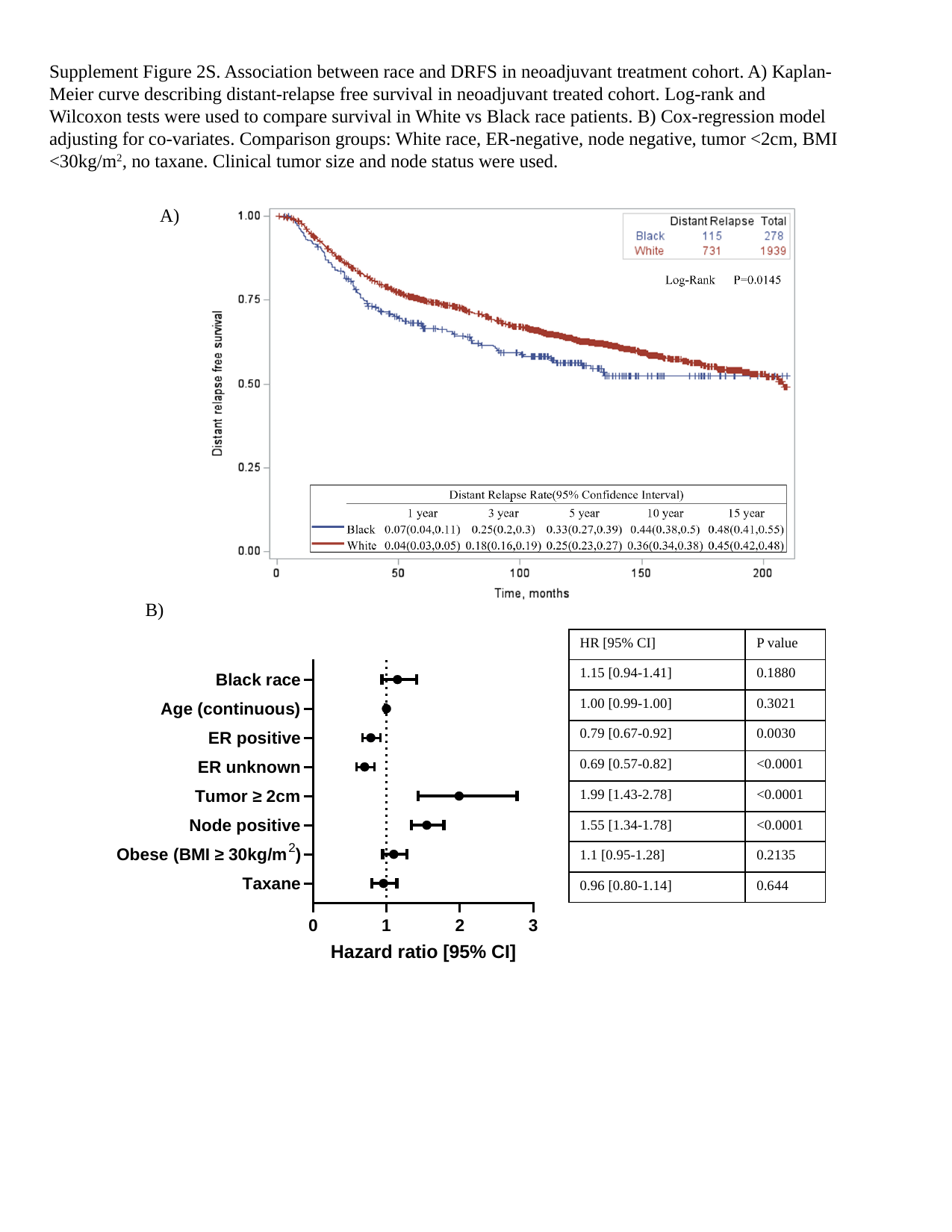

Supplement Figure 2S. Association between race and DRFS in neoadjuvant treatment cohort. A) Kaplan-Meier curve describing distant-relapse free survival in neoadjuvant treated cohort. Log-rank and Wilcoxon tests were used to compare survival in White vs Black race patients. B) Cox-regression model adjusting for co-variates. Comparison groups: White race, ER-negative, node negative, tumor <2cm, BMI <30kg/m^2^, no taxane. Clinical tumor size and node status were used.
A)
B)
| HR [95% CI] | P value |
| --- | --- |
| 1.15 [0.94-1.41] | 0.1880 |
| 1.00 [0.99-1.00] | 0.3021 |
| 0.79 [0.67-0.92] | 0.0030 |
| 0.69 [0.57-0.82] | <0.0001 |
| 1.99 [1.43-2.78] | <0.0001 |
| 1.55 [1.34-1.78] | <0.0001 |
| 1.1 [0.95-1.28] | 0.2135 |
| 0.96 [0.80-1.14] | 0.644 |

### Slide 11
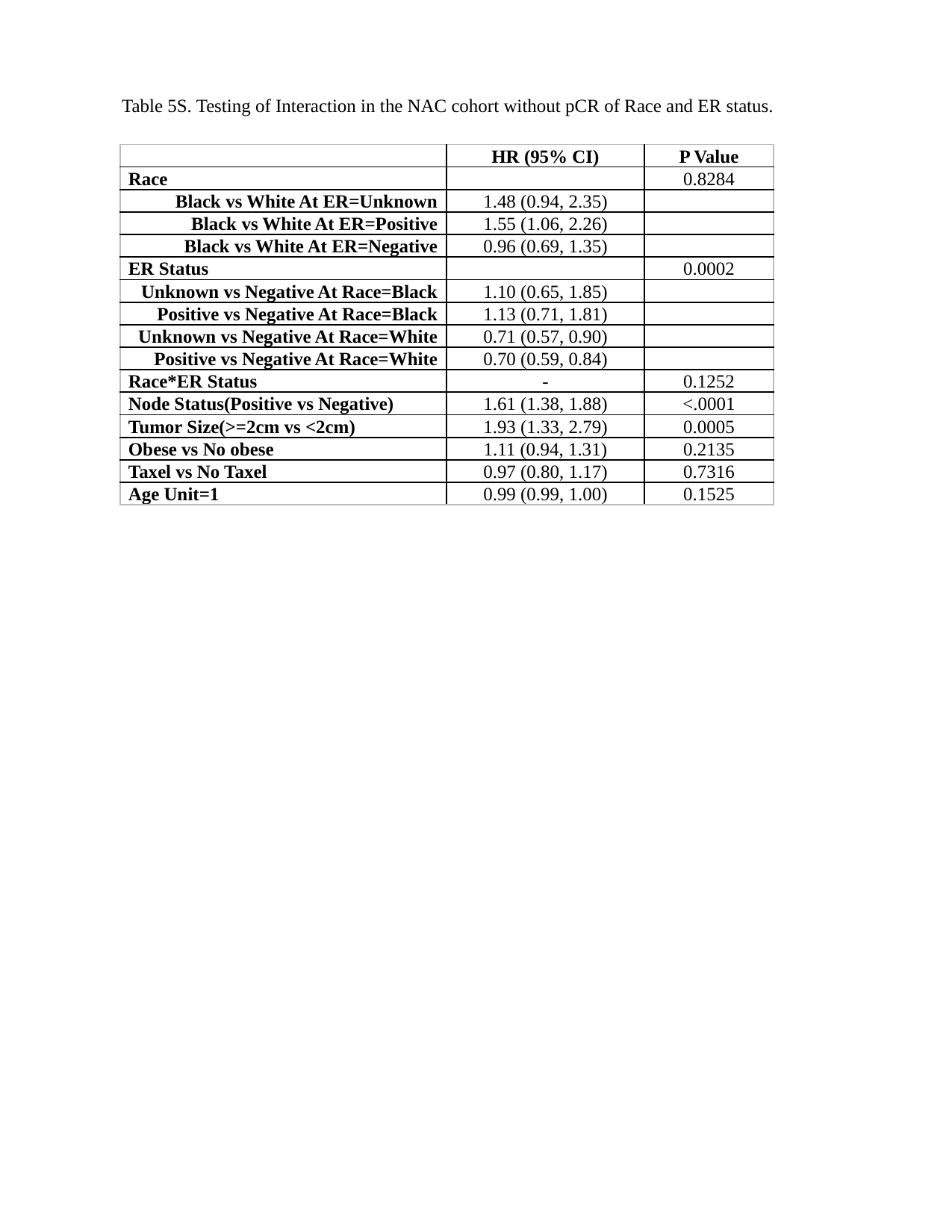

Table 5S. Testing of Interaction in the NAC cohort without pCR of Race and ER status.
| | HR (95% CI) | P Value |
| --- | --- | --- |
| Race | | 0.8284 |
| Black vs White At ER=Unknown | 1.48 (0.94, 2.35) | |
| Black vs White At ER=Positive | 1.55 (1.06, 2.26) | |
| Black vs White At ER=Negative | 0.96 (0.69, 1.35) | |
| ER Status | | 0.0002 |
| Unknown vs Negative At Race=Black | 1.10 (0.65, 1.85) | |
| Positive vs Negative At Race=Black | 1.13 (0.71, 1.81) | |
| Unknown vs Negative At Race=White | 0.71 (0.57, 0.90) | |
| Positive vs Negative At Race=White | 0.70 (0.59, 0.84) | |
| Race\*ER Status | - | 0.1252 |
| Node Status(Positive vs Negative) | 1.61 (1.38, 1.88) | <.0001 |
| Tumor Size(>=2cm vs <2cm) | 1.93 (1.33, 2.79) | 0.0005 |
| Obese vs No obese | 1.11 (0.94, 1.31) | 0.2135 |
| Taxel vs No Taxel | 0.97 (0.80, 1.17) | 0.7316 |
| Age Unit=1 | 0.99 (0.99, 1.00) | 0.1525 |
